# Anti-cancer effect of memantine as adjunctive therapy in metastatic colon cancer: A pilot randomized controlled clinical trial

**DOI:** 10.1101/2024.09.01.24312896

**Authors:** Kosar Jannesar, Yousef Roosta, Naser Masoudi, Rahim Asghari, Javad Rasouli, Hamid Soraya

**Author notes:** **Corresponding author** Dr. Hamid Soraya Department of Pharmacology, Faculty of Pharmacy, Urmia University of Medical Sciences, Urmia-Iran PO Box: 571571441.

## Abstract

**Purpose:** Colon cancer, one of the three deadliest cancers worldwide, has a high prevalence, especially in developing societies. Recently, our preclinical study demonstrated the strong anti-tumor effects of memantine on colon cancer in rats. This study aimed to investigate the effects of memantine (an NMDA receptor antagonist) in patients with metastatic colon cancer.

**Patients and Methods:** In this randomized controlled clinical trial, 32 patients with metastatic colon cancer were randomized into two arms. The first arm received a chemotherapy regimen and the second arm received a chemotherapy regimen plus memantine 20 mg/day. The tumor size, metastasis, hematological parameters, CEA level, and N/L ratio were measured. Additionally, we assessed the safety and tolerability of this combination and its effect on the quality of life (QoL) of metastatic colon cancer patients.

**Results:** Memantine reduced the colon tumor size in comparison to the control group patients (P=0.04). Also, in the memantine group, the metastasis was lower than in the control group (50% vs 87.5% respectively). Moreover, the memantine-treated group demonstrated reduced levels of CEA (P=0.01) as well as improved some hematological parameters. Also, quality of life was partially improved and no serious adverse effects were reported.

**Conclusions:** Three-month adjuvant therapy with memantine reduces tumor size, metastasis, CEA level, and the N/L ratio, and also causes relative improvement of hematological parameters as well as the quality of life without causing any serious adverse effects. Therefore, memantine could be suggested as an appropriate adjuvant therapy in metastatic colorectal cancer.

## Introduction

Colon cancer, one of the three deadliest cancers worldwide, has a high prevalence, especially in developing societies (1). Studies have envisioned that the onset of this malignancy comes from the genetic and epigenetic mutations of epithelial cells of the colon which leads to an increase in proliferation and occurrence of benign adenomas (2). Various types of risk factors have been recognized in progression of this disease. Familial hereditary, obesity, low level of physical activity, smoking and other life style related characteristics can be considered as the most prevalent risk factors (2, 3). Along with physical examination (the first step of diagnosis), screening methods such as colonoscopy, fecal blood test and sigmoidoscopy are the most important diagnostic procedures of metastatic colon cancer (4, 5). From the beginning of the diagnosis of colon cancer, various guidelines have been designed for the treatment of this malignancy, most of which include the use of chemotherapy drugs or surgery (6). The dominant chemotherapy regimen is based on the fluoropyrimidine class of drugs, with irinotecan and oxaliplatin as adjuvants. FOLFIRI (5-fluorouracil+oxaliplatin) and FOLIFOX (5-fluorouracil+irinotecan) as two prominent combination chemotherapy regimens showed progression in the reduction of metastasis and brings improvement for colon cancer patients. However, due to adverse effects such as nephrotoxicity, alopecia, diarrhea, etc., the treatment process can be disrupted and the patient’s quality of life can be affected. So, newer studies focused researches on combination of these regimens with targeted therapies such as bevascizumab (Anti-VEGF monoclonal antibody that restricts angiogenesis) (6–8). Despite many advances in the field of metastatic colon cancer treatment, the mortality rate is still high. Therefore, it seems necessary to search for agents with high efficacy and low adverse effects.

One of the important factors in diagnosis and monitoring of treatment is the level of biomarkers related to cancer. This can be very useful in evaluating the stage of malignancy, disease prognosis and treatment process. (9). One of the most important of these biomarkers is carcinoembryonic antigen (CEA), which is a non-specific biomarker that can be measured in the serum of patients (10). Beside of this biomarker, the level of neutrophil-to-lymphocyte ratio (N/L ratio) as one of the main inflammation biomarkers is elevated in colon cancer malignancy and lymph node spread (11). As an affordable method, this ratio can be a valuable guide in the process of colon cancer treatment. It has been shown that high levels of N/L ratio contributed significantly with the decrement in overall survival of colon cancer patients (12).

Glutamate as one of the vital neurotransmitters of CNS, is linked to the pathophysiology of different diseases. The N-Methyl-D-aspartate receptor (NMDAR) is a glutamate ion channel that is expressed increasingly in various cancers such as colon cancer and prostate malignancy (13, 14). The release of glutamate is associated significantly with proliferation and uncontrolled growth of gastric cancer cells and it has been proved that the inhibition of NMDA receptors can converse this process which indicates the importance of these receptors in diagnosis and treatment processes of cancer (14, 15). Memantine, as a NMDA receptor inhibitor, was initially used in the treatment of Alzheimer’s disease, but later, due to its effects on various biomarkers, researchers began to investigate its effects on different types of cardiovascular, neurological and cancer diseases (16). In 2016, Motagi et al showed that memantine could attenuates the levels of IL-6, MPO and TNF-α that leads to a reduction in ulcerative colitis (17). Also, memantine through inhibition of NMDA receptors causes a significant decrement in the survival of breast cancer cells and even induces NMDA-R mediated autophagic death in malignant glioma cells (18, 19). Treatment of prostate cancer cells with memantine decreased survival of cancer cells that is associated with the reduction of expression of oncogene c-Myc (20). Recently, our research team showed the anti-tumor effects of memantine in DMH-induced colon cancer in rats (21). However, the effects of memantine in patients with metastatic colon cancer are not known. In this randomized controlled trial, we intend to evaluate the efficacy of memantine as an adjuvant therapy to chemotherapy regimen in metastatic colon cancer patients. The results show a strong anticancer effect in metastatic colon cancer.

## Material and Methods

### Design and study sample

The present study was conducted as a pilot randomized controlled trial (RCT). In this study, patients with metastatic colorectal cancer referred to the blood and oncology clinic of Urmia Imam Khomeini Hospital, who were newly diagnosed and to start treatment, after obtaining informed consent, were randomly included in the study. They were divided into two equal groups of 16 patients with the treatment regimen of standard drugs plus memantine (memantine group) and the group receiving standard drugs (control group). The randomization process was carried out under the supervision of an epidemiologist using the block randomization method according to the corresponding sample size (Figure 1). This RCT was conducted under the supervision of the Ethics Committee of Urmia University of Medical Sciences (IR.UMSU.REC.1400.302). This study was registered in Iranian Registry of Clinical Trials (IRCT registration number: IRCT20210819052230N1)

**Figure 1.**
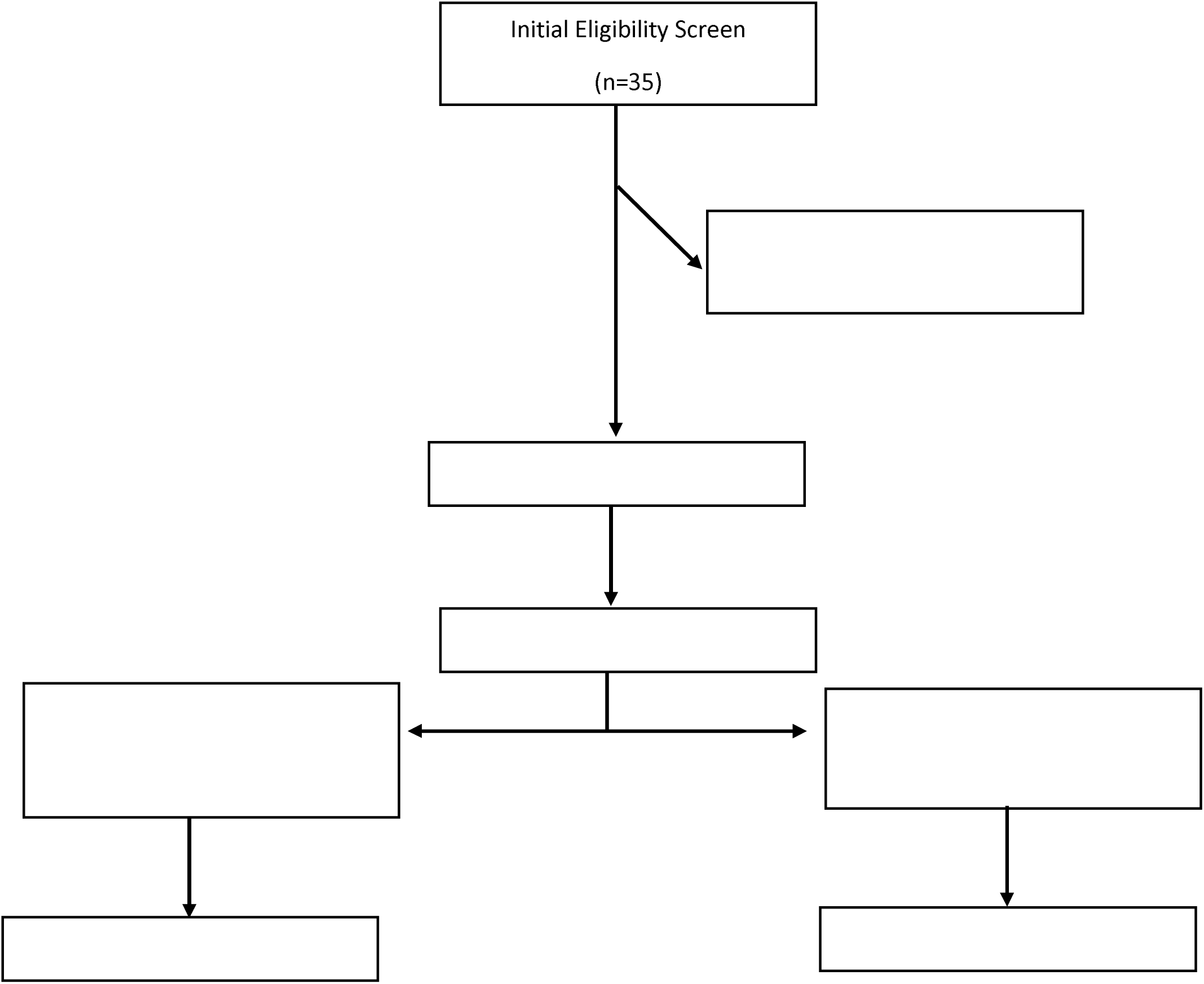
Randomized Trial Diagram.

### Eligibility criteria

#### Inclusion criteria include

All patients over 18 years of age with metastatic colon cancer who do not have a severe renal failure (15-29 CLcr (min/mL), liver failure (more than 5-fold increase in liver enzymes), and history of sensitivity to memantine, history of seizures, cardiovascular diseases, eye diseases, and simultaneous use of drugs that alkalize the pH of urine.

#### Exclusion criteria include

Drug sensitivity to memantine and intolerance to memantine during treatment, chronic alcohol consumption, addiction and abuse of drugs that reduce the level of consciousness such as benzodiazepines, decreased GFR, incidence of hepatotoxicity, gastrointestinal complications and patient non-cooperation (22).

### Intervention

The study was randomized and according to the usual procedure for all patients in the clinic, the file was created and the information related to age, sex, CT scan report of the patient and concurrent diseases and preliminary tests were included in the file. In all patients, standard chemotherapy protocol and other conventional treatments were implemented unchanged. In the patients of the memantine group, in addition to the standard chemotherapy regimen, memantine (Tasnim pharmaceutical industry, Iran) was given at a dose of 20 mg per day for 3 months (starting with a dose of 5 mg per day and an additional 5 mg per week and a maximum dose of 20 mg per day) and the other group was given the standard chemotherapy regimen without memantine (control group). At the beginning of the study, the conditions of the study were explained to the patient, and after obtaining informed consent, memantine was prescribed. The standard treatment regimen in the oncology department is based on NRAS and KRAS mutations, and the regimens used include; Bavacizumab + FOLFOX and Bevacizumab + FOLFIRI. It should be noted that while emphasizing the correct and timely use of prescription drugs, patients were requested to contact their physician immediately if they see any side effects or changes in the treatment process.

### Measures

Before starting the treatment (day 0), all the tests which are necessary to start chemotherapy and CEA level (by ELISA method) were requested by the specialist doctor and the data were entered in the checklists that were designed for this purpose. Also, the size and metastasis rate of tumors were measured using CT scan. At the end of the third month (day 90), the data in two groups were compared in terms of changes in body mass index, changes in N/L ratio, CBC and CEA level, the size of the primary tumor and its metastasis. It should be noted that during the study, two blood samples were taken from the patients, one at the beginning of the treatment and the other at the end of the third month (day 90), both of which were due to routine follow-up tests by a specialist.

### Assessment of health-related quality of life

The EORTC QLQ-C30 questionnaire was used to assess the quality of life of colon cancer patients. This questionnaire comprises a 30-item that includes a global Quality of Life (QoL) scale, 5 functional scales (physical, role, emotional, cognitive, and social functioning), 3 multi-item symptom scales (fatigue, nausea/vomiting, and pain), and 6 single-item symptom scales (dyspnea, sleep disturbance, appetite loss, constipation, diarrhea, and financial difficulties) (23). All participants underwent two interviews; the initial interview occurred at baseline, coinciding with the first chemotherapy cycle (day 0), while the subsequent interview took place three months later (day 90). Subsequently, the EORTC QLQ-C30 underwent scoring in accordance with the guidelines outlined in the EORTC scoring manual (24). Following the scoring process, responses to Likert scale questions were transformed into a 0–100 scale in a linear manner. Higher scores on functional scales and the global scale are indicative of enhanced Quality of Life, whereas heightened scores on symptom scales and individual items suggest increased severity of symptoms and diminished Quality of Life.

### Statistical analysis

For quantitative data, central and dispersion indices (mean and standard deviation), and for qualitative variables, frequency and percentage of frequency were calculated and statistical tables and graphs were used to display the data as needed. To compare the two groups according to the modality and data distribution (normality), parametric statistical tests such as paired t-test or independent t-test equivalents were used. The significance level for all tests was considered less than 0.05 (p<0.05). All analyzes were performed using Graph pad prism 9 software.

## Results

In this randomized controlled trail, we had two groups of control and memantine 20 mg/day. In control group, the patients with metastatic colon cancer received chemotherapy according to the standard protocol of our hospital. In memantine group, the patients with metastatic colon cancer received chemotherapy according to the standard protocol plus memantine 20 mg per day for 3 months (90 days). We had 16 patients in each groups and also we had 16 males and 16 female patients. The age of the patients in the control group was 45.19±3.39 and in the memantine treated group was 52.19±2.8 which the difference was not significant. Regarding the body mass index (BMI) of studied patients also there was no significant difference between two groups. Please see table 1.

**Table 1.** Gender, age and BMI of patients.

|  | **Control (n=16)** | **Memantine (n=16)** | **P value** |
| --- | --- | --- | --- |
| Gender | 9 female<br>7 male | 9 male<br>7 female | - |
| Age | 45.19±3.39 | 52.19±2.8 | 0.12 |
| BMI day 0 | 23.25±1 | 22.63±0.97 | 0.67 |
| BMI day 90 | 21.31±0.78 | 20.81±0.79 | 0.65 |

### The effects of memantine on hematological parameters

The serum analysis of patients demonstrated that at day 0 (start of study) the white blood cells (WBC) count in the control group was 22413±14037 and in the memantine treated group was 70681±18544 which is significant difference (P=0.046), but three months later and at the end of study (day 90) there was no significant difference between control and memantine groups. HCT at day 0 in the control group was 34.00±1.09 and in the memantine group was 28.28±1.11 which shows significant difference (P=0.001), but after 90 days’ treatment there was no significant difference between groups. For detailed information please see table 2.

**Table 2.** Hematological parameters in the control and memantine treated groups’ patients. All data are presented as the mean±SEM and an unpaired t-test was used to compare the two control and memantine groups. WBC: White blood cells; HGB: Hemoglobin; HCT: Hematocrit; MCV: Mean corpuscular volume; MCH: Mean corpuscular hemoglobin; MCHC: Mean corpuscular hemoglobin concentration; PLT: Platelet count.

|  | Control day 0 | Memantine day 0 | P value | Control day 90 | Memantine day 90 | P value |
| --- | --- | --- | --- | --- | --- | --- |
| WBC | 22413±14037 | 70681±18544 | 0.046* | 14713±7715 | 11956±6652 | 0.78 |
| HGB | 11.74±0.29 | 11.58±0.29 | 0.71 | 11.56±0.3 | 11.93±0.28 | 0.37 |
| HCT | 34.00±1.09 | 28.28±1.11 | 0.001* | 32.23±1.67 | 30.12±1.13 | 0.3 |
| MCV | 86.08±2.22 | 71.81±3.23 | 0.001* | 79.23±3.17 | 63.03±4.34 | 0.005* |
| MCH | 29.22±0.81 | 29.34±1.07 | 0.92 | 27.09±1.05 | 30.61±1.24 | 0.03* |
| MCHC | 33.76±0.3892 | 34.03±0.43 | 0.68 | 32.86±0.46 | 33.07±0.52 | 0.77 |
| PLT | 185625±24816 | 225625±12535 | 0.16 | 189563±19951 | 221188±17571 | 0.24 |
| Neutrophil | 5438±1018 | 9438±603.9 | 0.002* | 1390±150.2 | 1274±89.15 | 0.51 |
| Lymphocyte | 2144±243.8 | 1688±146.3 | 0.11 | 1861±174.7 | 1675±138.3 | 0.4 |

### The effects of memantine on tumor size and metastasis

Our results showed that there was no significant difference in tumor size between control (25.6±1.9 mm) and memantine groups (25.8±2.68 mm) at day 0 (start of study). Three months’ treatment with memantine 20 mg once a day reduced tumor size from 23.7±1.88 in the control group to 17.44±2.23 mm in the memantine treated group (P=0.04). Moreover, unlike the control group, comparing the tumor size in the memantine group at day 0 and day 90 (pre-post analysis) demonstrated a significant effect (P=0.0008) on tumor size (Figure 2).

**Figure 2.**
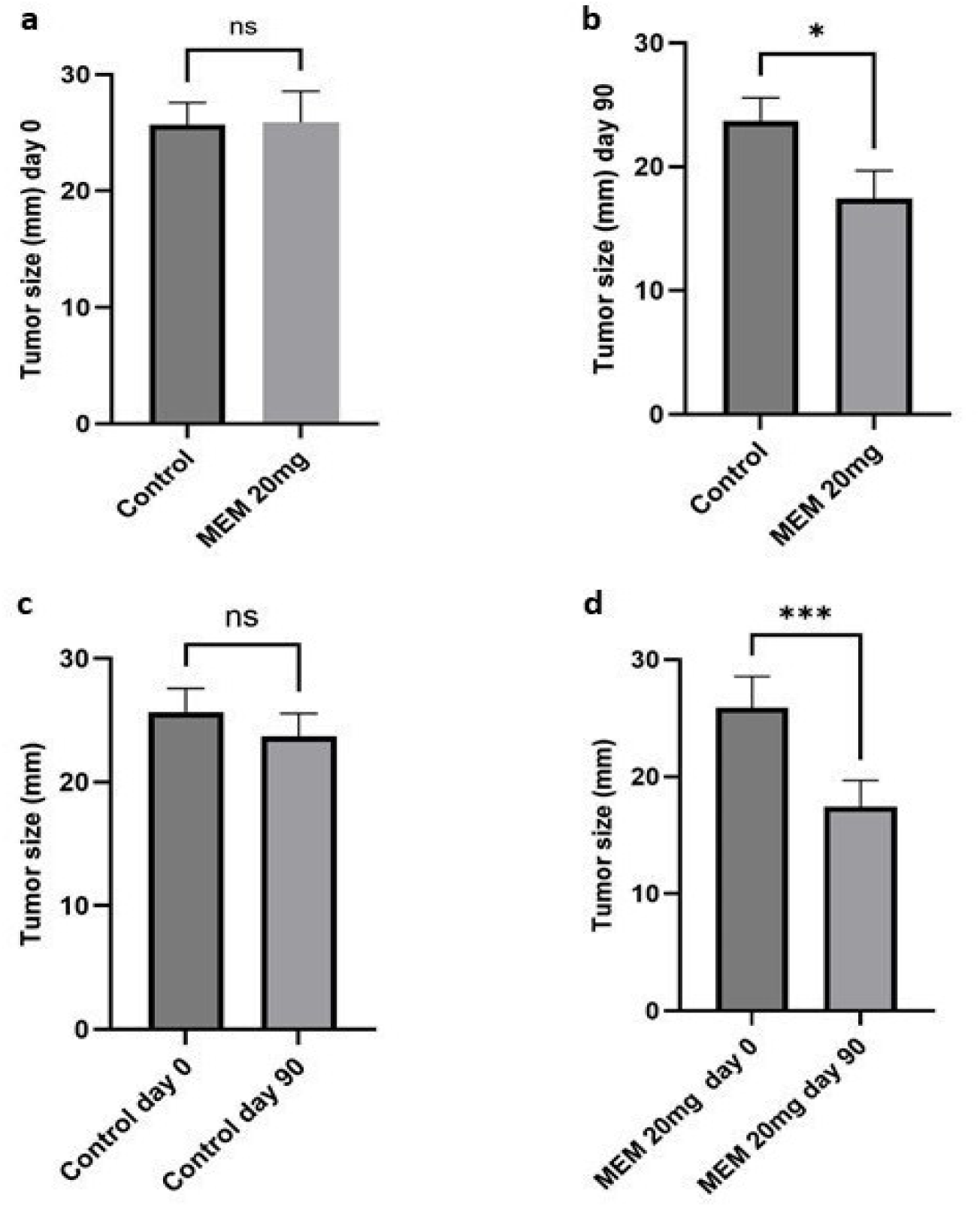
The effect of memantine on tumor size in metastatic colon cancer patients. a) Tumor size in the control and memantine groups at day 0 (start of study). b) Tumor size in the control and memantine groups at day 90 (end of study). c) Comparison of tumor size in the control group at day 0 and day 90. d) Comparison of tumor size in the memantine group at day 0 and day 90. All data are expressed as the mean±SEM and an unpaired t-test was used to compare control and memantine groups in *a* and *b*. In *c* and *d* for comparison in each group at day 0 and day 90 (pre-post) we used paired t-test. P< 0.05 was considered statistically significant. *P=0.04; ***P=0.0008

All of the studied patients had metastatic colon cancer which was diagnosed and confirmed by our research team oncologist. So, all patients in both studied groups had metastasis at day 0 but surprisingly, three months’ treatment with memantine 20 mg per day demonstrated a very valuable effect and reduced colon cancer metastasis. We observed that in the control group 2 patients (12.5%) were free of metastasis, but in the memantine treated group 8 patients (50%) were free of metastasis at the end of the study (day 90) (Figure 3).

**Figure 3.**
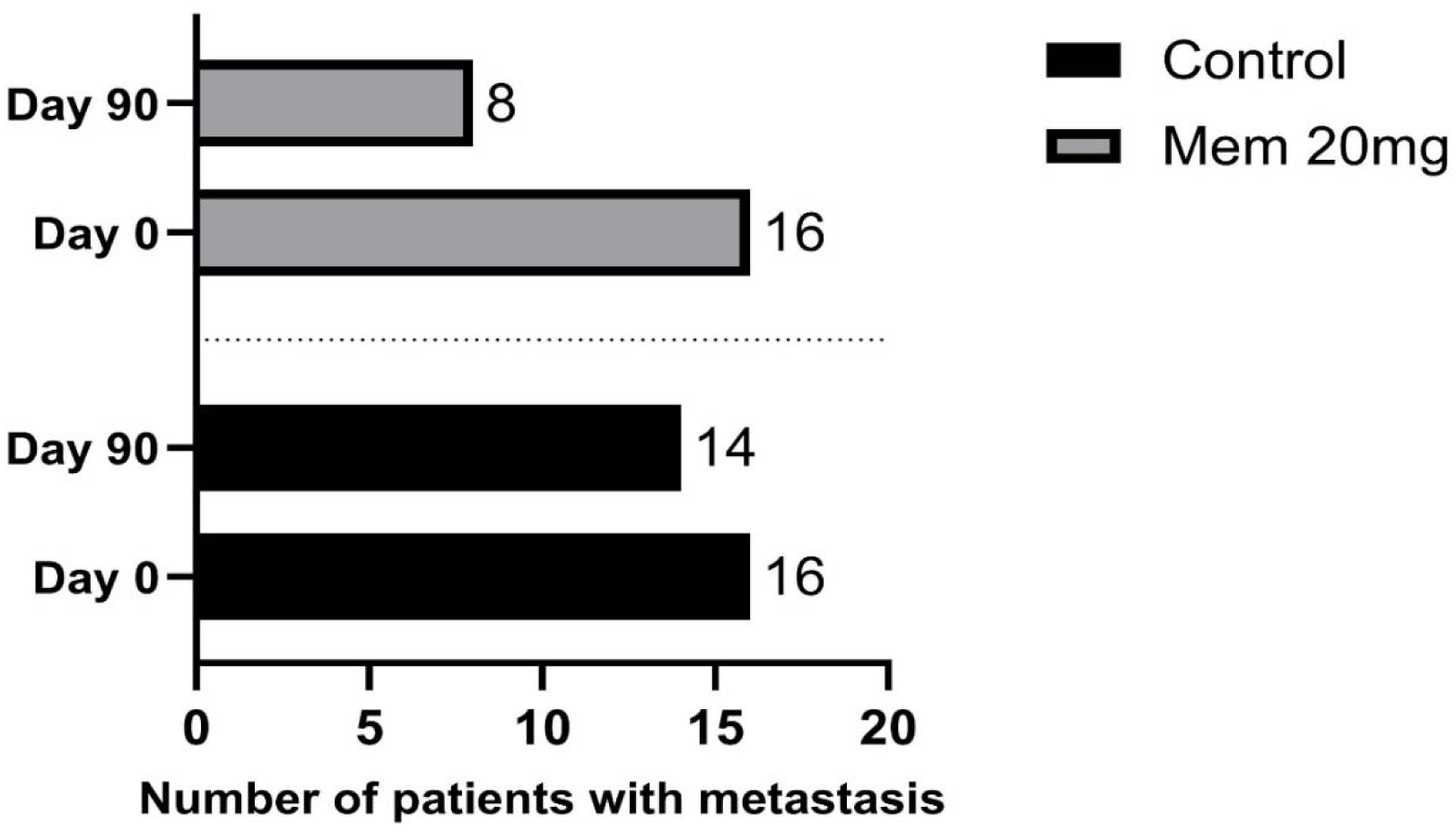
The effect of memantine on metastasis in metastatic colon cancer patients.

### The effects of memantine on serum CEA levels

The CEA levels at day 0 was similar between control (88.59±6.84 ng/mL) and memantine treated (85.29±5.18 ng/mL) groups. Three months’ treatment with memantine 20 mg once a day reduced significantly the CEA levels from 47.4±9.3 ng/mL in the control group to 15.93±5.23 (P=0.01). Further analysis inside each group showed that treatment with standard chemotherapy protocol (control group) reduced the level of CEA by 46.5% and adding memantine to standard chemotherapy protocol (memantine group) decreased the level of CEA by 81.3% which was a profound effect (Figure 4).

**Figure 4.**
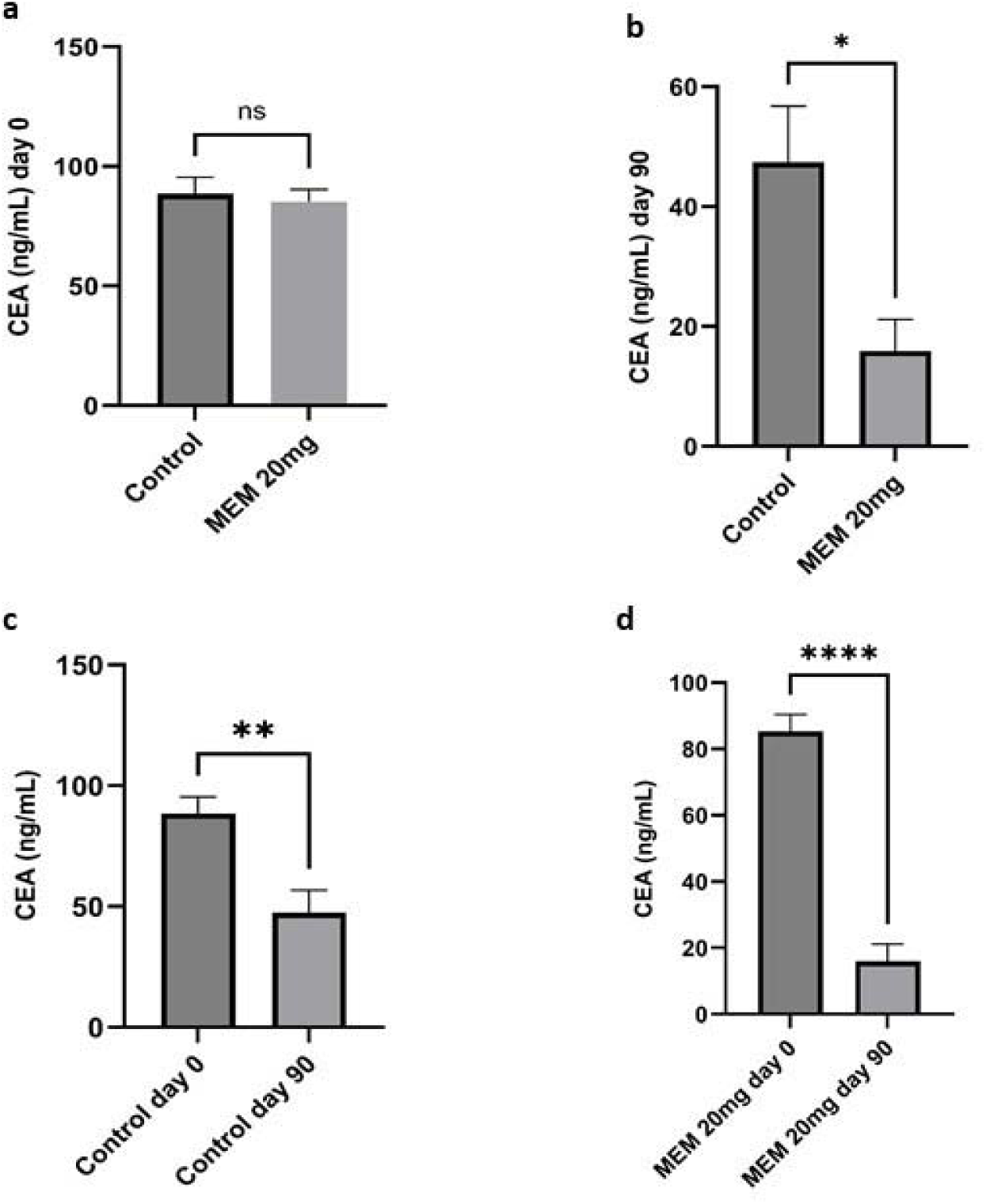
The effect of memantine on CEA levels in metastatic colon cancer patients. a) CEA levels in the control and memantine groups at day 0 (start of study). b) CEA levels in the control and memantine groups at day 90 (end of study). c) Comparison of CEA levels in the control group at day 0 and day 90. d) Comparison CEA levels in the memantine group at day 0 and day 90. All data are expressed as the mean±SEM and an unpaired t-test was used to compare control and memantine groups in *a* and *b*. In *c* and *d* for comparison in each group at day 0 and day 90 (pre-post) we used paired t-test. *P=0.01; **P=0.001; ****P<0.0001

### The effects of memantine on neutrophil to lymphocyte ratio

The neutrophil-to-lymphocyte ratio (N/L ratio), which is calculated as a ratio between the neutrophil and lymphocyte counts, is an important prognostic factor for metastatic colon cancer. Our results showed that N/L ratio at day 0 was different between control (2.67±0.31) and memantine (6.24±0.67) groups (P<0.0001). Treatment in both groups reduced the N/L ratio to the same level. Further analysis in each group (pre-post analysis) demonstrated that treatment with standard chemotherapy protocol (control group) reduced the N/L ratio by 69% and adding memantine to standard chemotherapy protocol (memantine group) decreased the N/L ratio by 203% which was a profound effect (Figure 5).

**Figure 5.**
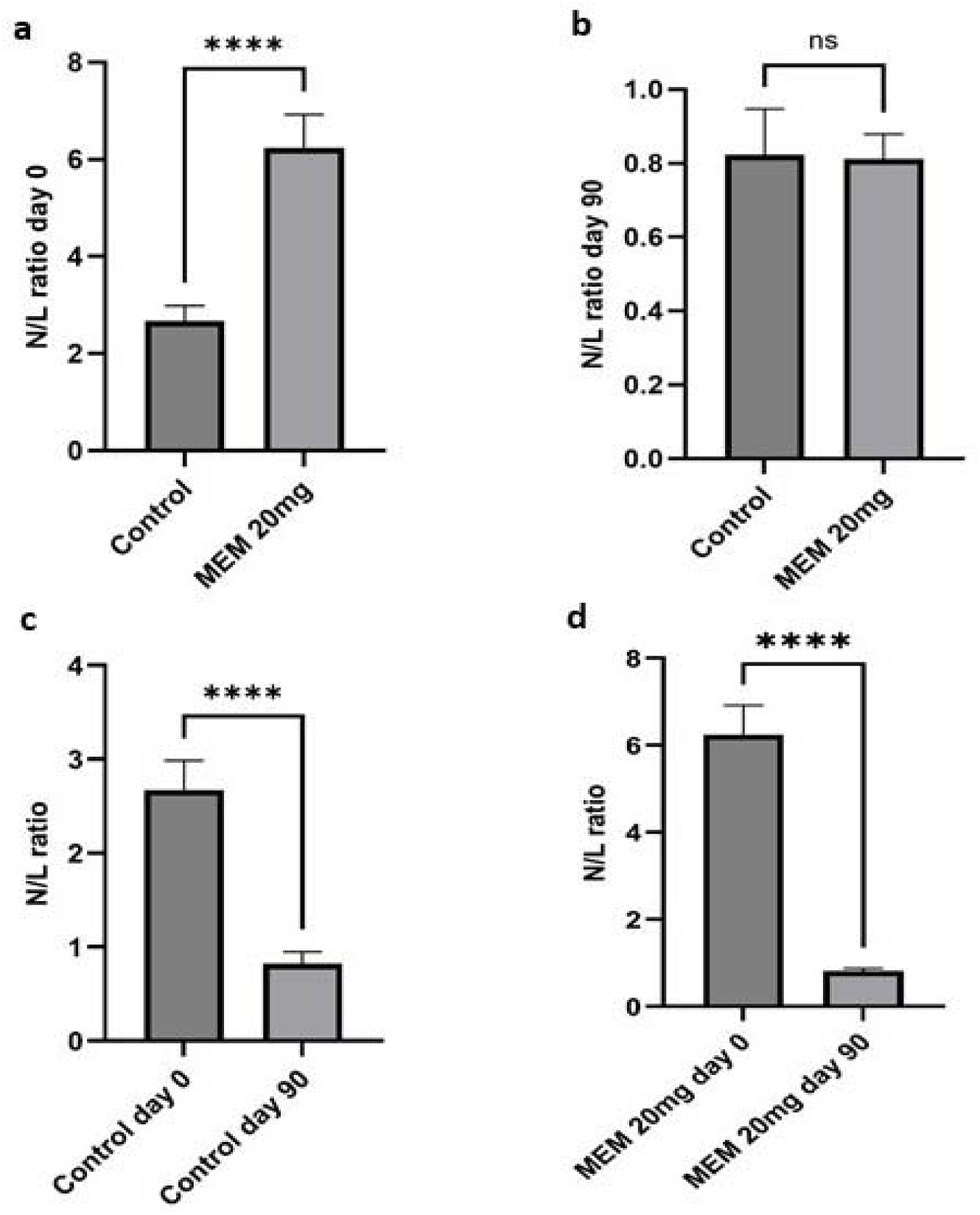
The effect of memantine on neutrophil to lymphocyte (N/L) ratio in metastatic colon cancer patients. a) N/L ratio in the control and memantine groups at day 0 (start of study). b) N/L ratio in the control and memantine groups at day 90 (end of study). c) Comparison of N/L ratio in the control group at day 0 and day 90. d) Comparison of N/L ratio in the memantine group at day 0 and day 90. All data are expressed as the mean±SEM and an unpaired t-test was used to compare control and memantine groups in *a* and *b*. In *c* and *d* for comparison in each group at day 0 and day 90 (pre-post) we used paired t-test. ****P<0.0001.

### The effects of memantine on health-related quality of life

Using the EORTC QLQ-C30, baseline mean scores for functional scales, symptoms scales, and global quality of life, were almost similar and there were no significant differences between the control group and the memantine-treated group (Table 3). After three months of treatment, the memantine group demonstrated better cognitive (P=0.01) and role functioning (P=0.04) in comparison to the control group. Moreover, memantine-treated patients showed lower constipation (P=0.03). All other scales were not significantly different between the two studied groups (Table 4).

**Table 3.** Comparison of the mean scores of EORTC QLQ-C30 global quality of life, functional, and symptoms scales between the control group and the memantine group at baseline (day 0).

| Variables | Baseline mean scores ± SEM | | P-value |
| --- | --- | --- | --- |
|  | Control group | Memantine group |  |
| Functional Scales and Global QoL |  |  |  |
| Physical Functioning | 34.7±5.8 | 36.8±6 | 0.8 |
| Role Functioning | 13±5.8 | 12.9±4.6 | 0.99 |
| Emotional Functioning | 16.6±5.5 | 18.8±5.4 | 0.77 |
| Cognitive Functioning | 40.5±7.9 | 54.1±9.6 | 0.28 |
| Social Functioning | 7.1±3.7 | 15.2±5.4 | 0.22 |
| Global Health | 65.5±4 | 64±3.9 | 0.78 |
| Symptom Scales |  |  |  |
| Fatigue | 93.6±3 | 88±3.7 | 0.26 |
| Nausea and Vomiting | 74.9±9.5 | 70.2±9.4 | 0.72 |
| Pain | 94.4±2.6 | 89.2±3.7 | 0.26 |
| Dyspnea | 45.2±10.8 | 38±9.1 | 0.61 |
| Insomnia | 66.6±9.2 | 62.2±10.7 | 0.75 |
| Appetite Loss | 82±7.1 | 76.8±6.9 | 0.61 |
| Constipation | 57.1±10.1 | 40.4±8.6 | 0.22 |
| Diarrhea | 56.3±10.2 | 46.1±12.2 | 0.52 |
| Financial Difficulties | 69±9.5 | 49.9±10.3 | 0.18 |

**Table 4.**
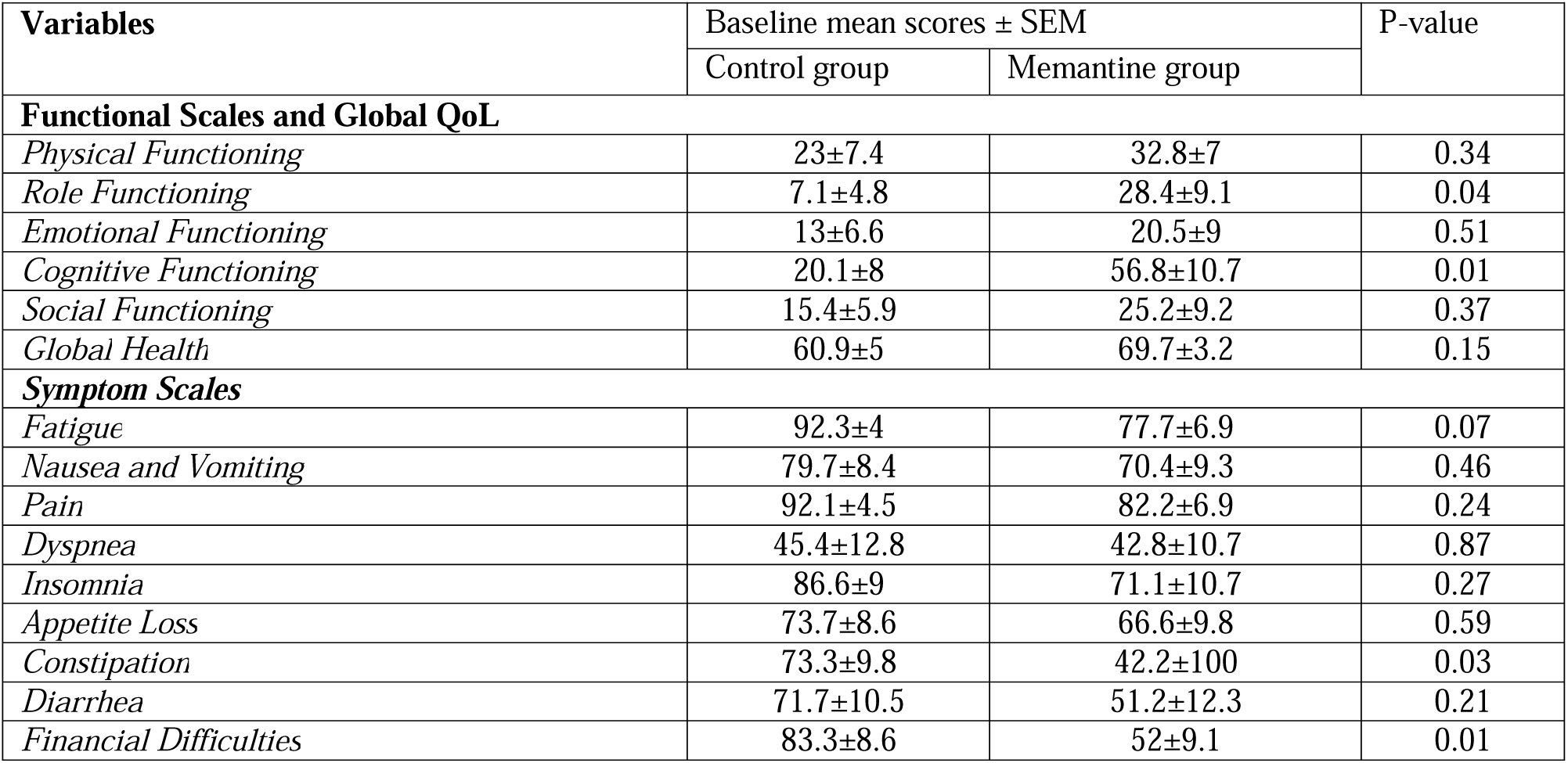
Comparison of the mean scores of EORTC QLQ-C30 global quality of life, functional, and symptoms scales between the control group and the memantine group at the end of study (day 90).

## Discussion

As to the fact that metastatic colon cancer is one of the most lethal cancers in the world and because of the lack of an effective and safe treatment for this malignancy, in this study we aimed to investigate the effects of memantine (an NMDA receptor antagonist) on patients with metastatic colon cancer.

We have demonstrated that three months of adjuvant therapy with memantine (20 mg/day) along with chemotherapy regimen surprisingly reduced the colon tumor size in comparison to control group patients. Also, a brilliant result that we reach to, is that, in the memantine group 50% of patients were free of metastasis in comparison to 12.5% of control group after 90 days’ treatment. Moreover, it was observed that the level of CEA was lowered by 81.3% in memantine treatment group regard to the first day of study that could be considered as a better result in comparison to 46.5% decrease of CEA in control group from day 0 to day 90. Measuring N/L ratio as one of the prognostic factors, also revealed that, the decrement of this ratio in memantin group was a notable percent of 203% whereas this reduction in control group was just 69%. Furthermore, evaluation of hematological parameters revealed that consumption of memantine caused several positive effects. WBC, MCH and HCT levels in the treatment group patients became closer to the normal range of parameters at the end of the study in comparison to control group and measuring of neutrophile levels showed that memantine was able to reduce the level of these cells on the contrary of control group.

Along with pathophysiological roles of NMDA receptors in CNS, these glutamate receptors have shown various potential effects in different tissues. Also, evident is the fact that NMDA receptors are overexpressed in cancer cell lines and cause an increase in the invasive growth of tumors and even their migration. It has been observed that the inhibition of NMDA receptors (genetically or pharmacologically) leads to a reduction in the growth and metastasis of tumor cells (13). One of the NMDA receptor antagonists named AP5, has shown anti-proliferative effects in gastric cancer cells (25). Moreover, studies has demonstrated that memantine and dizocilpine (MK-801) has a potential effect of the controlling of various cancer cell lines growth and metastasis including lung, prostate, and pancreatic cells (26–28). Genetically knock down of NMDA receptors in breast to brain metastasis, considered as an effective therapeutic procedure for significant reduction of tumor infiltration (29). Furthermore, Ketamine as a NMDA receptor antagonist demonstrated outstanding effects in colon cancer and by a reduction in VEGF level and tumor metastasis, this agent attenuates tumor cell migration that leads to an important success in the treatment of metastatic colon cancer (30). In this trial, along with previous studies, it has been shown that, prescription of memantine for three months besides of standard chemotherapy regimen, brings a significant decrease in terms of tumor size and metastasis.

Carcinoembryonic antigen (CEA) is one of the main serum biomarkers which has a prognostic importance in the treatment period. The level of this biomarker is increased in various malignancies such as colon cancer, breast cancer and ovarian cancer (31–33). Normal level of CEA is usually reported in the range of 0-2.9 ng/ml and it has been proven that the higher levels of 5 ng/ml indicates colorectal cancer and levels that are greater than 35 ng/ml are associated with cancer reoccurrence. In colorectal cancer higher levels of CEA is associated with the severity of malignancy and also it has been confirmed that this biomarker could be considered as a prominent detector for restaging and recurrence of colorectal cancer even in post-surgery cases (34, 35). In evaluation of newer therapies for colon cancer, measurement of this biomarker level before starting the novel treatment and after 3 months therapy could be a vital factor in assessment of effects of new procedure (36, 37). Evaluation of CEA levels after 8 weeks of chemotherapy in metastatic colorectal cancer patients, confirmed the fact that this biomarker reduction by 50% is a helpful indicator for the treatment success (38). A clinical trial by Cho et al. on the immunomodulatory effects of ketamine in colorectal cancer patients showed that, intraoperative use of ketamine (0.25 mg/kg before surgery and an infusion of 0.05 mg/kg/hour) has no significant changes on the level of inflammatory responses and CEA level after two years after surgery (39). However, in this study, our results demonstrated that memantine at the dosage of 20mg/day for 3 months significantly reduced the CEA level and so can be an effective agent in controlling of tumor recurrence.

Neutrophil/Lymphocyte ratio as one of the important inflammatory biomarkers, is usually calculated in different malignancies including colorectal cancer and has a prognostic importance in evaluation of therapies (40). Prediction of patient’s survival with measurement of this ratio, and also the simplicity and cheapness of it, makes it a good choice for assessment of treatment (41). Furthermore, the calculating of this ratio can give some information regarding to overall survival of colorectal cancer patients along with the possibility of disease relapse (42). Higher levels of this ratio is contributed significantly to the stage 4 of colorectal cancer that altered us for high risk of disease recurrence. So, a decrease in this ratio means to a better survival and lower risk of metastasis (43). It has been observed that, memantine and MN-08 (a novel nitrate derivative of memantine) significantly decreased N/L ratio in sepsis-induced lung injury mice and so, it could be considered as a novel therapy (44). Our results, likewise, confirmed the results of previous studies and showed that, memantine significantly reduced the N/L ratio by 203% in day 90 in comparison to day 0, whereas, in control group the changes were just a low of 69% in the end of the study. This shows the strong effect of memantine and supports its use in metastatic colon cancer.

Several previous studies have demonstrated that hematological parameters could be considered as valid factors in the evaluation of the effects of treatments in colorectal cancer. Duo to difficulty and high costs of other examinations, viewing the levels of hematological parameters and comparing them in the onset and end of the study is a valuable strategy (45–47). The evaluation of CBC results demonstrated that, contrary to the beginning of the study, where there was a significant difference in the amount of WBC level between two groups (p=0.46), the use of memantine in patients brought the level of this parameter closer to the normal amount. Also, it has been shown that, memantine consumption leads to an increase in the level of HCT whereas, in the control group, the amount of this parameter was lower at the end of study. These changes abolish the significant differences of two groups (p=0.001 at the onset of study and p=0.3 at the end) and revealed that memantine help patients to have a normal range of HCT level (35–50). Memantine has the same effects on MCH levels whereas its usage caused a minor increase in the amount of this parameter (unlike the control group) and this leads to a significant difference between two groups at the end of the study (P=0.03). These alternations in the MCH levels, meanwhile aid patients to experience the very closer levels to normal range of MCH. The most striking change in the levels of hematological parameters is related to neutrophil levels, where, the significant difference at the first day of study (P=0.002) conversed to an insignificant one (P=0.51) due to the impressive reduction of neutrophil changes in treatment group in comparison to control one. The mentioned changes in hematological parameters are all in the direction of the positive effects of memantine in improving the condition of patients compared to the control group and can be considered as one of the positive therapeutic features of this drug.

Moreover, in order to support the beneficial effects of memantine in patients with metastatic colon cancer, we can mention the improving effects of memantine in the quality of life of these patients. Treatment with memantine improved some quality of life factors in the EORTC QLQ-C30 questionnaire such as better cognitive and role functioning as well as constipation in comparison to the control group. In other words, not only were no serious side effects observed with memantine, but it also caused a relative improvement in the patient’s quality of life.

In conclusion, this pilot randomized controlled clinical trial showed that a three-month administration of memantine at a dose of 20 mg daily along with a chemotherapy regimen can reduce tumor size, metastasis, CEA level, and the N/L ratio, and also cause relative improvement of hematological parameters as well as quality of life without causing any serious adverse effects. Therefore, memantine could be suggested as an appropriate adjuvant drug with a chemotherapy regimen in metastatic colorectal cancer.

## Data Availability

All data produced in the present study are available upon reasonable request to the authors
All data produced in the present work are contained in the manuscript

## Author contributions

**K. Jannesar:** Investigation, data curation, methodology, writing–original draft. **Y. Roosta:** Data curation, methodology, supervision, validation, editing. **N. Masoudi:** Conceptualization, methodology, visualization, validation, review and editing. **R. Asghari:** Conceptualization, methodology, validation, review and editing. **J. Rasouli:** Formal analysis, methodology, validation, review and editing. **H. Soraya:** Conceptualization, data curation, formal analysis, supervision, methodology, visualization, validation, writing–review and editing.

## Data Availability Statement

All data generated or analyzed during this study are included in this published article and further datasets are available from the corresponding author on reasonable request.

## References

1. Rawla P, Sunkara T, Barsouk A. Epidemiology of colorectal cancer: incidence, mortality, survival, and risk factors. Gastroenterology Review/Przegląd Gastroenterologiczny. 2019;14(2):89–103.

2. Hossain MS, Karuniawati H, Jairoun AA, Urbi Z, Ooi DJ, John A, et al. Colorectal cancer: a review of carcinogenesis, global epidemiology, current challenges, risk factors, preventive and treatment strategies. Cancers. 2022;14(7):1732.

3. Low EE, Demb J, Liu L, Earles A, Bustamante R, Williams CD, et al. Risk factors for early-onset colorectal cancer. Gastroenterology. 2020;159(2):492–501. e7.

4. Ahmed M. Colon cancer: a clinician’s perspective in 2019. Gastroenterology research. 2020;13(1):1.

5. Cappell MS. The pathophysiology, clinical presentation, and diagnosis of colon cancer and adenomatous polyps. Medical Clinics. 2005;89(1):1–42.

6. Sanz-Garcia E, Grasselli J, Argiles G, Elez ME, Tabernero J. Current and advancing treatments for metastatic colorectal cancer. Expert opinion on biological therapy. 2016;16(1):93–110.

7. Gómez-España M, Gallego J, González-Flores E, Maurel J, Páez D, Sastre J, et al. SEOM clinical guidelines for diagnosis and treatment of metastatic colorectal cancer (2018). Clinical and Translational Oncology. 2019;21:46–54.

8. Biller LH, Schrag D. Diagnosis and treatment of metastatic colorectal cancer: a review. Jama. 2021;325(7):669–85.

9. Srivastava S, Verma M, Henson DE. Biomarkers for early detection of colon cancer. Clinical Cancer Research. 2001;7(5):1118–26.

10. Gugoasa LA, Stefan-van Staden R-I, Al-Ogaidi AJM, Stanciu-Gavan C, van Staden JF, Rosu M-C, et al. Molecular recognition of colon cancer biomarkers: P53, KRAS and CEA in whole blood samples. Journal of the Electrochemical Society. 2017;164(9):B443.

11. Xia L-j, Li W, Zhai J-c, Yan C-w, Chen J-b, Yang H. Significance of neutrophil-to-lymphocyte ratio, platelet-to-lymphocyte ratio, lymphocyte-to-monocyte ratio and prognostic nutritional index for predicting clinical outcomes in T1–2 rectal cancer. BMC cancer. 2020;20:1–11.

12. Naszai M, Kurjan A, Maughan TS. The prognostic utility of pre-treatment neutrophil-to-lymphocyte-ratio (NLR) in colorectal cancer: a systematic review and meta-analysis. Cancer Medicine. 2021;10(17):5983–97.

13. Gallo S, Vitacolonna A, Crepaldi T. NMDA receptor and its emerging role in cancer. International Journal of Molecular Sciences. 2023;24(3):2540.

14. Stepulak A, Rola R, Polberg K, Ikonomidou C. Glutamate and its receptors in cancer. Journal of neural transmission. 2014;121:933–44.

15. Seidlitz EP, Sharma MK, Saikali Z, Ghert M, Singh G. Cancer cell lines release glutamate into the extracellular environment. Clinical & experimental metastasis. 2009;26:781–7.

16. Shafiei-Irannejad V, Abbaszadeh S, Janssen PM, Soraya H. Memantine and its benefits for cancer, cardiovascular and neurological disorders. European Journal of Pharmacology. 2021;910:174455.

17. Motaghi E, Hajhashemi V, Mahzouni P, Minaiyan M. The effect of memantine on trinitrobenzene sulfonic acid-induced ulcerative colitis in mice. European Journal of Pharmacology. 2016;793:28–34.

18. Seifabadi S, Vaseghi G, Javanmard SH, Omidi E, Tajadini M, Zarrin B. The cytotoxic effect of memantine and its effect on cytoskeletal proteins expression in metastatic breast cancer cell line. Iranian Journal of Basic Medical Sciences. 2017;20(1):41.

19. Yoon W-S, Yeom M-Y, Kang E-S, Chung Y-A, Chung D-S, Jeun S-S. Memantine induces NMDAR1-mediated autophagic cell death in malignant glioma cells. Journal of Korean Neurosurgical Society. 2017;60(2):130.

20. Albayrak G, Konac E, Dikmen A, Bilen C. Memantine induces apoptosis and inhibits cell cycle progression in LNCaP prostate cancer cells. Human & experimental toxicology. 2018;37(9):953–8.

21. Jannesar K, Eftekhari P, Pourjabali M, Masoudi N, Soraya H. Anti-tumor effect of memantine, an N-methyl-D-aspartate receptor antagonist, against DMH-induced colon cancer in rats. Journal of research in pharmacy (online). 2022;26(2):345–53.

22. Cheong CK, Nistala KRY, Ng CH, Syn N, Chang HSY, Sundar R, et al. Neoadjuvant therapy in locally advanced colon cancer: a meta-analysis and systematic review. J Gastrointest Oncol. 2020;11(5):847–57.

23. Barakat HE, Hussein RR, Elberry AA, Zaki MA, Ramadan ME. The impact of metformin use on the outcomes of locally advanced breast cancer patients receiving neoadjuvant chemotherapy: An open-labelled randomized controlled trial. Scientific Reports. 2022;12(1):7656.

24. Fayers P, Aaronson N, Bjordal K, Groenvold M, Curran D, Bottomley A. EORTC QLQ-C30 scoring manual the EORTC QLQ-C30 introduction. EORTC QLQ-C30 Scoring Man. 2001;30:1–67.

25. Watanabe K, Kanno T, Oshima T, Miwa H, Tashiro C, Nishizaki T. The NMDA receptor NR2A subunit regulates proliferation of MKN45 human gastric cancer cells. Biochemical and biophysical research communications. 2008;367(2):487–90.

26. North WG, Gao G, Jensen A, Memoli VA, Du J. NMDA receptors are expressed by small-cell lung cancer and are potential targets for effective treatment. Clinical pharmacology: advances and applications. 2010:31–40.

27. Abdul M, Hoosein N. N-methyl-D-aspartate receptor in human prostate cancer. The Journal of membrane biology. 2005;205:125–8.

28. Li L, Zeng Q, Bhutkar A, Galván JA, Karamitopoulou E, Noordermeer D, et al. GKAP acts as a genetic modulator of NMDAR signaling to govern invasive tumor growth. Cancer Cell. 2018;33(4):736–51. e5.

29. Zeng Q, Michael IP, Zhang P, Saghafinia S, Knott G, Jiao W, et al. Synaptic proximity enables NMDAR signalling to promote brain metastasis. Nature. 2019;573(7775):526–31.

30. Duan W, Hu J, Liu Y. Ketamine inhibits colorectal cancer cells malignant potential via blockage of NMDA receptor. Experimental and Molecular Pathology. 2019;107:171–8.

31. Téllez-Avila FI, García-Osogobio SM. [The carcinoembryonic antigen: apropos of an old friend]. Rev Invest Clin. 2005;57(6):814–9.

32. Morimoto Y, Takahashi H, Arita A, Itakura H, Fujii M, Sekido Y, et al. High postoperative carcinoembryonic antigen as an indicator of highlZIrisk stage II colon cancer. Oncology letters. 2022;23(5):1–8.

33. Tao C, Rouhi J. A biosensor based on graphene oxide nanocomposite for determination of carcinoembryonic antigen in colorectal cancer biomarker. Environmental Research. 2023;238:117113.

34. Konishi T, Shimada Y, Hsu M, Tufts L, Jimenez-Rodriguez R, Cercek A, et al. Association of Preoperative and Postoperative Serum Carcinoembryonic Antigen and Colon Cancer Outcome. JAMA Oncol. 2018;4(3):309–15.

35. Litvak A, Cercek A, Segal N, Reidy-Lagunes D, Stadler ZK, Yaeger RD, et al. False-positive elevations of carcinoembryonic antigen in patients with a history of resected colorectal cancer. J Natl Compr Canc Netw. 2014;12(6):907–13.

36. Locker GY, Hamilton S, Harris J, Jessup JM, Kemeny N, Macdonald JS, et al. ASCO 2006 update of recommendations for the use of tumor markers in gastrointestinal cancer. J Clin Oncol. 2006;24(33):5313–27.

37. Kankanala VL, Mukkamalla SKR. Carcinoembryonic antigen. 2022.

38. Colloca GA, Venturino A, Guarneri D. Carcinoembryonic antigen-related tumor kinetics after eight weeks of chemotherapy is independently associated with overall survival in patients with metastatic colorectal cancer. Clinical Colorectal Cancer. 2020;19(4):e200–e7.

39. Sun CJ, Young KN, Jae-Kwang S, Hae JJ, Lee S, Young-Lan K. The immunomodulatory effect of ketamine in colorectal cancer surgery: a randomized-controlled trial. Canadian Journal of Anesthesia. 2021;68(5):683–92.

40. Pine J, Morris E, Hutchins G, West N, Jayne D, Quirke P, et al. Systemic neutrophil-to-lymphocyte ratio in colorectal cancer: the relationship to patient survival, tumour biology and local lymphocytic response to tumour. British journal of cancer. 2015;113(2):204–11.

41. Gao S, Tang W, Zuo B, Mulvihill L, Yu J, Yu Y. The predictive value of neutrophil-to-lymphocyte ratio for overall survival and pathological complete response in breast cancer patients receiving neoadjuvant chemotherapy. Front Oncol. 2022;12:1065606.

42. Mazaki J, Katsumata K, Kasahara K, Tago T, Wada T, Kuwabara H, et al. Neutrophil-to-lymphocyte ratio is a prognostic factor for colon cancer: a propensity score analysis. BMC Cancer. 2020;20(1):922.

43. Tsai P-L, Su W-J, Leung W-H, Lai C-T, Liu C-K. Neutrophil–lymphocyte ratio and CEA level as prognostic and predictive factors in colorectal cancer: A systematic review and meta-analysis. Journal of cancer research and therapeutics. 2016;12(2):582–9.

44. Hu H, Jiang H, Zhang K, Zhang Z, Wang Y, Yi P, et al. Memantine nitrate MN-08 suppresses NLRP3 inflammasome activation to protect against sepsis-induced acute lung injury in mice. Biomedicine & Pharmacotherapy. 2022;156:113804.

45. Karacan E, Yilmaz EM, Yildiz B, Demirkiran AE. The contribution of hematological parameters to prediction of the phase in stage 2 and stage 3 colon cancers. Journal of Clinical Medicine of Kazakhstan. 2020(3 (57)):35-8.

46. Hu Z, Tan S, Chen S, Qin S, Chen H, Qin S, et al. Diagnostic value of hematological parameters platelet to lymphocyte ratio and hemoglobin to platelet ratio in patients with colon cancer. Clinica Chimica Acta. 2020;501:48–52.

47. Dere Ö, Dere Y. The Impact of Hematologic Parameters on Histopathologic Features of Colorectal Cancer. International Journal of General Medicine. 2024:2029–36.

